# Exploring the impact of a context-adapted decision aid and online training about shared decision making about goals of care with elderly patients in the intensive care unit: a mixed-methods study

**DOI:** 10.1101/2024.09.07.24313154

**Authors:** Ariane Plaisance, Julien Turgeon, Lucas Gomes Souza, France Légaré CQ, Stéphane Turcotte, Nathalie Germain, Tommy Jean, Maude Dionne, Félix Antoine Fortier, Patrick Plante, Diane Tapp, Véronique Gélinas, Emmanuelle Bélanger, Mark H Ebell, Christian Chabot, Tom van de Belt, Alexis F Turgeon, Patrick M Archambault

## Abstract

**Purpose:** To explore the impact of a context-adapted decision aid and an online training about shared decision-making (SDM) about goals of care on the level of involvement of elderly patients by intensivists in SDM about goals of care and quality of goals of care discussions (GCD) in an intensive care unit.

**Methods:** This was a three-phase before-after mixed-methods implementation study conducted in an ICU in Lévis, Quebec, Canada. We followed the StaRI and COREQ reporting guidelines. We recruited patients aged ≥ 65 and their attending intensivists. We video-recorded GCD in three phases: Phase I: GCD without a decision aid; Phase II: GCD with a decision aid about goals of care but no online training; and Phase III: GCD with both a decision aid about goals of care following online training about SDM. All GCD recordings were transcribed verbatim. We measured the level of patient engagement by intensivists in SDM about goals of care through the OPTION scale and evaluated GCD quality using the Audit of Communication, Care Planning, and Documentation (ACCEPT) indicators. A qualitative thematic analysis of the encounters transcriptions was also performed.

**Results:** Out of 359 eligible patients, the study included 21 patients (71% males; median age, 77 years; 57% without high school diploma) and 5 intensivists (80% male; median age, 35). Despite completing online training, the decision aid was never used in recorded encounters. We did not perform any tests of statistical significance to compare results in each study phase because of small sample sizes over each phase. OPTION and ACCEPT scores were low in each phase, but physicians did engage in GCD. We found that 76% of the goals of care recorded in medical records after the discussion were consistent with preferences expressed by patients during recorded observations. Several patients expressed confusion about GCD. Barriers identified by intensivists leading GCD include physician attitudes, challenges to performing GCD along with the demands of the intensive care unit, misunderstandings, and lack of training. Facilitators include a patient-centered approach, a clear decision aid, and positive patient attitudes. In future work, an environment that supports physicians in performing GCD, promotes earlier and higher quality patient GCD before admission to the intensive care unit, and encourages meaningful SDM in critical care must be assessed as pathways to successful intensive care unit GCD.

**Conclusion:** A context-adapted decision aid about goals of care was created in addition to a complementary online training module. The online training was completed by all participating physicians but no increased involvement of patients in SDM during intensive care unit GCD was observed, and use of the decision aid was also not observed. We found several communication barriers that will need to be explored to improve intensive care unit GCD.

**Trial registration number:** NCT04034979

**Key points:** Misunderstandings and concerns among older adult patients about goals of care and invasive interventions in the ICU contribute to delayed decision-making.

An online training regarding shared decision making with a corresponding decision aid for discussing goals of care was completed by all participating intensivists, but no increased involvement of patients nor use of the decision aid was observed in the ICU.

Facilitators to the uptake of shared decision making may include the involvement of non-intensivist health professionals, mandating documentation discussions and their results in patient files, and challenging a long-held reluctance to discuss death as an outcome.

## 1 Introduction

Older adults near the end of life often receive life-sustaining interventions that may not be congruent with their preferences nor their goals of care [1]. Even though most older people value comfort and quality of life over prolongation of life by life-sustaining therapies [2–4], their medical orders do not reflect those preferences in 70% of cases [5]. Goals of care discussions (GCD) with hospitalized elderly patients can increase the congruence between patients’ preferences and their medical care [6,7]. During GCD, patients and clinicians discuss patients’ prognosis, level of functional autonomy, values and life goals in order to inform decisions regarding the potential use of life-sustaining interventions [8]. Clinical practice guidelines recommend shared decision making (SDM) to facilitate these discussions [8,9]. SDM involves patients and clinicians making a decision together based on the best available evidence, clinician’s experience and expertise and what matters to the patients [10,11]. Decision aids (DAs) are structured evidence-based tools aiming to provide unbiased information and guidance to support SDM with patients facing difficult health decisions [12,13]. Consequently, health professionals trained in SDM are more likely to use DAs and adopt SDM with patients than professionals not trained in SDM [10]. From the patient’s point of view, the adoption of GCD, SDM, and DAs may represent a meaningful shift towards more personalized and value-driven care at the end of life [14,15]. This shift means a transition from passive recipients of care to active and respected partners in the decision-making process, ensuring that their end of life decision-making is aligned with their personal values and desires.

We developed a DA supporting GCD adapted to the local context of a single intensive care unit (ICU) in the province of Québec, Canada employing a user-centered design [16]. We explore the impact of using the context-adapted DA as the intervention under study coupled with an online training program about and designed to implement shared decision-making (SDM) in an intensive care unit (ICU). We observed and analyzed the subsequent level of elderly patient involvement in SDM by intensivists during goals of care discussions (GCD) and quality of GCD in the ICU. Our hypothesis was that using the context-adapted DA supported by training in SDM would increase clinicians’ involvement of elderly patients in SDM about goals of care and the quality of GCD.

## 2 Methods

### 2.1 Design

This was a three-phase before-after mixed methods implementation project [17]. In Phase I (2017-05-22 to 2017-07-23), we recorded patient-intensivist discussions on GCD without a DA. In Phase II (2017-07-25 to 2017-09-28), we recorded GCD with a DA option available for intensivists, but no training. In Phase III (2017-11-06 to 2018-01-25), we recorded another set of GCD, allowing intensivists to use the DA after completing SDM training. The study was reported using the Standards for Reporting Implementation Studies (StaRI) [18] and the Consolidated criteria for reporting qualitative research (COREQ) [19]. The project was approved by the Centre intégré de santé et de services sociaux de Chaudière-Appalaches (CISSS-CA) Research Ethics Board (Certificate #2017-001).

### 2.2 Setting

The monocentric study took place in a medical and surgical ICU (18 beds and six intensivists) of a university teaching hospital in Lévis, Canada. The research team was led by a female PhD student in community health sciences (AP) and the principal investigator (PA), a male intensivist at the ICU site and clinician scientist. Both are experienced in qualitative studies and mixed methods. Two female research coordinators at the CISSSCA, each holding an MSc (MD, VG) and a male internal medicine resident doing rotations at this ICU (JT) were also involved with discussions, interviews, data collection, and qualitative analysis.

### 2.3 Participants

Eligible patients had to be: a) aged ≥ 65 and b) capable of making their own healthcare decisions as determined by the attending intensivist’s clinical judgment. We excluded patients that were intubated, facing immediate urgent decisions, under the care of the principal investigator (PA), patients who did not read and speak French, and patients for whom the attending intensivist determined that a GCD was not necessary based on clinical judgment. All intensivists working in the ICU were eligible, but we excluded the principal investigator, fellows, residents and trainees.

### 2.4 Development of the decision aid and online training

Using the Knowledge to Action Framework [17], we developed a DA tool and an online training program that would be used during the intervention phases. We adapted and combined two existing DAs [20–22] to the local ICU context using user-centered design [16]. Our development strategy also included an analysis of the goals of care communication barriers with critically ill patients, their families, intensivists and other allied health professionals.

The resulting DA presents two invasive life-saving procedures (cardiopulmonary resuscitation and invasive mechanical ventilation) and contains questions to help patients reflect upon their goals of care and about the acceptability of different potential outcomes. The DA is also associated with an online calculator (wikidecision.org) based on the Good Outcomes Following Attempted Resuscitation (GO-FAR) prediction rule [21] linked to visual icons generated by an online visual risk representation application (<u>iconarray.com</u>, Risk Science Center and Center for Bioethics and Social Sciences in Medicine, University of Michigan, Ann Arbor, USA) to present the chances of survival with a favorable neurological outcome following in-hospital resuscitation.

The online training program was completed on average within a 60-minute timeframe and was developed for intensivists aimed to improve communication skills and SDM knowledge, as well as explain the DA. The content of this program (See Appendix 1 in Supplement) [23] was based upon three existing training programs about GCD and was reviewed by 11 experts in SDM, intensive care and palliative care [24–27].

### 2.5 Study procedures

#### 2.5.1 Data collection

The research team screened the list of all consecutive admitted ICU patients searching for eligible patients during normal working hours from Monday to Friday. Patients admitted to the ICU off-hours were still screened and considered eligible if the attending intensivist had not yet conducted a GCD. After introduction by the attending intensivist, a member of the research team obtained written consent to participate from the patient. Family members wishing to participate also completed a consent form.

We collected patients and intensivists baseline socio-demographic characteristics and patients baseline official goals of care. We also calculated Charlson comorbidity indices using clinical data [28]. The discussions between patients and intensivists were audio- or video recorded (based on patient’s preference) using a GoPro Hero5 (San Mateo, CA, USA) and each encounter was then transcribed verbatim and reviewed by the authors. No intervention was delivered to intensivists during Phase I to determine the current decision-making process about goals of care. During Phase II, the intensivists were given access to the DA, but without training on its use.

During Phase III, intensivists were trained about SDM including how to use the DA.

After each online training session, we collected intensivists’ self-reported completion time, satisfaction with the different parts of the training, self-reported perceived change in confidence to perform SDM and confidence to use the DA. To verify understanding of the online content, the intensivists participated in a one-hour debriefing session (total of 3 sessions) with authors (AP, MD, and PA). We used a structured verification list to make sure all intensivists understood the 9 essential elements of SDM [29] (Appendix 2).

#### 2.5.2 Outcomes

The primary implementation outcome was the impact of the use of our context-adapted DA and SDM training program on the level of patient’s involvement in SDM measured by the OPTION scale. As secondary outcomes we measured GCD quality (ACCEPT quality indicators) and characteristics. We also explored: a) elderly patients’ most frequent misunderstandings and concerns, b) clinicians’ evaluation of the training, and c) clinicians’ opportunities to improve their SDM skills in GCD. Finally, we also sought to determine if our intervention would improve the congruence between written medical orders for life-sustaining therapies and patient’s expressed preferences. All outcomes are detailed in the following sections.

Concerning patient involvement in SDM, we used the validated French version of the OPTION scale to measure the level of patient involvement by a clinician during a clinical encounter from an observer’s viewpoint [30,31] (Appendix 3) about cardiopulmonary resuscitation (CPR) and mechanical ventilation decision-making. OPTION scores were determined for each of the three implementation phases. Two trained reviewers (AP and JT) analyzed the recordings independently, but were not blinded to the intervention phase. They both scored each OPTION scale item on a scale of 0 to 4 using a coding guide (Appendix 4) and converted their scores into percentages (a high score/percentage is desirable and indicates higher involvement).

Disagreements were resolved by consensus between AP and JT.

Concerning the quality of goals of care discussions, we used an adapted version of the Conceptual Framework for Improving End-of-life Communication and Decision-making [7]. This framework evaluates the quality of end-of-life communication and decision making using 34 Audit of Communication, Care Planning, and Documentation (ACCEPT) quality indicators [32]. The 34 ACCEPT quality indicators are separated into four categories: advance care planning (8 items), GCD (13 items), documentation (5 items) and organization/system aspects (8 items). We only kept the seven quality indicators that were not similar to an item of the OPTION scale (Appendix 5). Two authors (TJ and JT) analyzed the discussions and rated them independently using both the recordings and the verbatim transcripts. Disagreements were resolved by consensus between AP and JT. ACCEPT quality indicators were determined for each of the three implementation phases.

#### 2.5.3 Goals of care discussion characteristics

We measured the use of the DA, length of the discussion and counted the number of words pronounced in total, by the physician and by the patient using a word count function. We also documented the presence of family members. We also collected ICU and hospital lengths of stay, and we documented all official written medical orders for life-sustaining therapy in the patient’s chart after each of the GCD.

#### 2.5.4 The most frequent misunderstandings and concerns of older patients

We performed qualitative content thematic analysis to identify patients’ most frequent misunderstandings and concerns and clinicians’ opportunities to improve their SDM skills in GCD. A misunderstanding was defined as a patient’s lack of understanding about the concept of goals of care, the interventions or the risks and benefits of the interventions. A concern was defined as a matter that seemed to cause feelings of unease, uncertainty, or apprehension to the patient.

#### 2.5.5 Barriers and facilitators to the context-adapted DA and training program

In March 2019, after the study was completed, we conducted a feedback session with all participating intensivists to gather their thoughts about the context-adapted DA, the training program, and SDM. During this session, the principal investigator (PA) presented the results of the study and led a discussion with the participants to gather their suggestions, their barriers and facilitators to SDM and improvements to the context-adapted DA and training program. This session was audio-recorded and transcribed verbatim.

### 2.6 Sample size

Our sample size was determined by three fixed predetermined two-month recruitment periods, based on the availability of research personnel for data collection. Based on our previous user-centered design study [16] that recruited similar patients in the same ICU during the same time period, we expected to recruit 10 patients per period to explore the main implementation issues [33]. All eligible intensivists in the ICU (*n* = 5) were included in the study.

### 2.7 Data Analysis

#### 2.7.1 Quantitative data analysis

We used descriptive statistics for categorical data and median, interquartile range (IQR) for continuous data describing patient, intensivists and clinical encounters characteristics. We also used descriptive statistics to present results for the OPTION scale items and for the ACCEPT quality indicators over three phases. We did not perform any statistical tests to compare OPTION scores or presence of ACCEPT quality indicators between study phases because our sample size was too small at each study phase. Even if different patients participated at each phase, the highly correlated nature of the behaviors being measured for each intensivist at each phase, a larger sample size would have been required to account for this clustering and correlation in our data.

#### 2.7.2 Qualitative data analysis

Two authors (AP and MD) conducted a discourse analysis [34] of all patient-intensivist encounter transcriptions using the R package for Qualitative Data Analysis [35] (version 0.2-7). Intensivists’ opportunities to improve their GCD skills were classified based on the OPTION scale. The two authors conducted their analyses separately on the full data set, final themes were determined by consensus. Two other authors (PA, VG) independently completed the thematic analysis of the transcribed post-study intensivist feedback session. Final themes were also determined by consensus. Participants’ representative quotes were translated from French and presented in English in this article.

## 3 Results

### 3.1 Patients, intensivists and clinical encounters characteristics

We recruited 21 patients, 7 per phase (Figure 1). All eligible intensivists (n = 5) participated in the study. Among excluded patients, a majority (72.5%; 227/313) were excluded by attending intensivists who decided not to engage in GCD themselves frequently stating that GCD had already been discussed. The encounters were video recorded except for one that was only audio recorded. Patient and intensivist characteristics are presented in Tables 1 and 2.

**Fig. 1.**
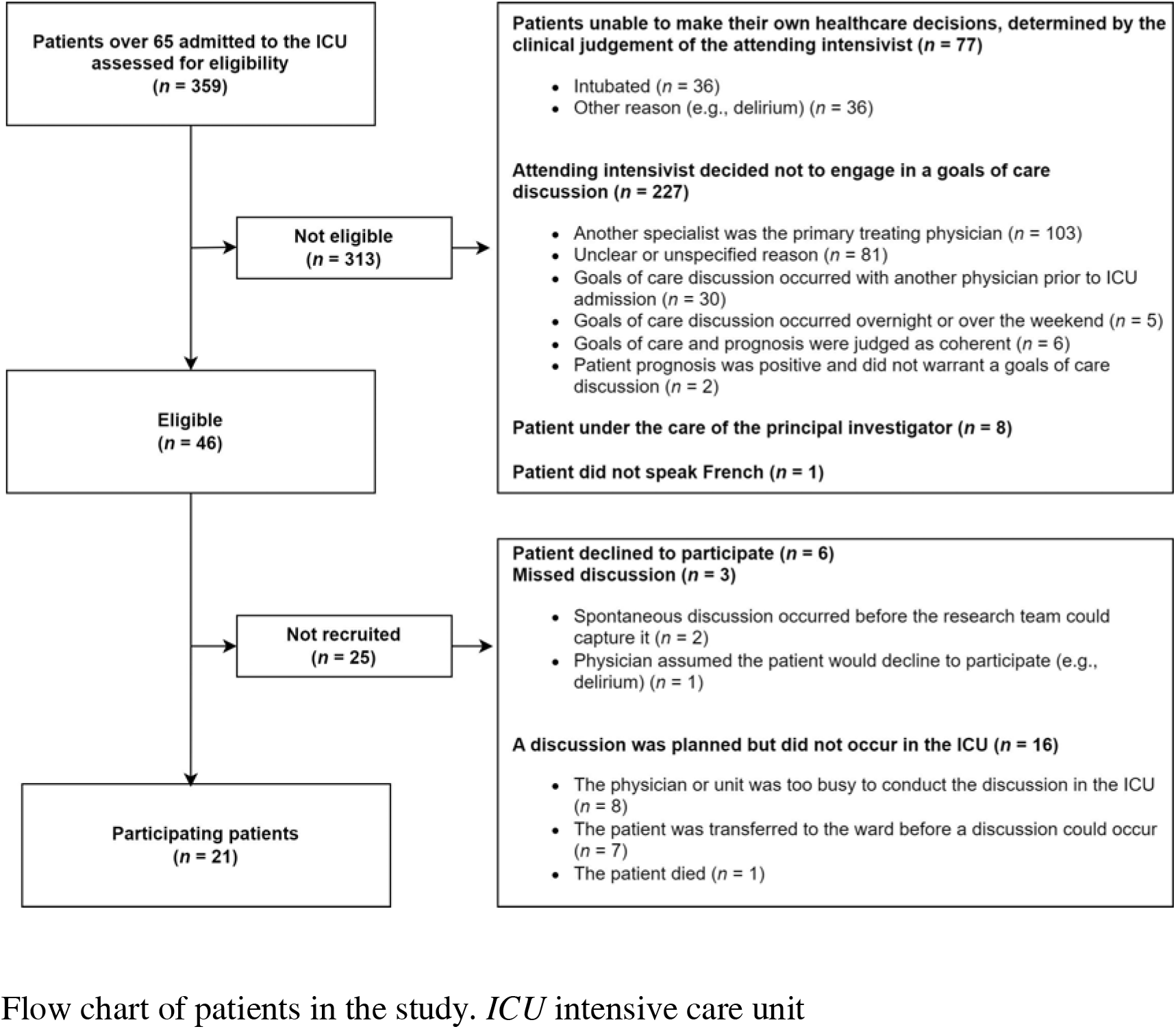
Flow chart of patients in the study. *ICU* intensive care unit.

**Table 1.** Patient baseline characteristics.

| **Patients** |  |  |  |  |
| --- | --- | --- | --- | --- |
|  | Phase 1<br>(*n* = 7) | Phase 2<br>(*n* = 7) | Phase 3<br>(*n* = 7) | Total<br>(*n* = 21) |
| **Men, *n* (%)** | 3 (42) | 6 (86) | 6 (86) | 15 (71) |
| **Age (years), median (IQR)** | 71 (70–84) | 80 (78–89) | 67 (65–79) | 77 (68–82) |
| **French Canadian, *n* (%)** | 6 (86) | 7 (100) | 7 (100) | 20 (95) |
| **Christian, *n* (%)** | 6 (86) | 7 (100) | 7 (100) | 20 (95) |
| **High school not completed** | 4 (58) | 5 (72) | 3 (42) | 12 (57) |
| **Married, *n* (%)** | 1 (14) | 5 (72) | 5 (72) | 11 (52) |
| **Charlson index, median (IQR)** | 5 (4–8) | 8 (5–12) | 8 (5–10) | 7 (5–10) |
| **Length of hospital stay (days), median (IQR)** | 6 (3–9) | 5 (4–30) | 10 (6–13) | 8 (4–15) |
| **Length of ICU stay (days), median (IQR)** | 3 (2–5) | 3 (2–8) | 5 (3–7) | 4 (3–6) |

*IQR* interquartile range, *n* count, *ICU* intensive care unit

**Table 2.** Intensivist sociodemographic characteristics.

| **Intensivists (*n* = 5)** |  |
| --- | --- |
| **Men, *n* (%)** | 4 (80) |
| **Age (years), median (IQR)** | 35 (33–43) |
| **Experience (years post residency), median (IQR)** | 4 (2–14) |
| **Fellowship in Critical Care, *n* (%)** | 5 (100) |

*IQR* interquartile range, *n* count

### 3.2 Involvement of patients in decision making by clinicians

Overall, we observed low OPTION scores in each phase and a small decrease in involvement scores across the three phases: 25% (Phase 1), 21% (Phase 2), and 19% (Phase 3). Due to a limited number of observed GCD at each phase (*n* = 7) (Table 3) and an unequal participation of intensivists at each phase with one intensivist contributing to 9 out of all the 21 discussions (Table 5), we were not able to perform any statistical test to compare clinicians’ involvement of patients in decision making at each phase.

**Table 3.** Clinician involvement in patient decision making measured by their OPTION scores and representative quotes of communication strategies used to engage patients.

| | Phase 1 ($n = 7$) | Phase 2 ($n = 7$) | Phase 3 ($n = 7$) |
| --- | --- | --- | --- |
| OPTION item and description | Median OPTION score (% [IQR]) | Median OPTION score (% [IQR]) | Median OPTION score (% [IQR]) |
| 1. The clinician draws attention to an identified problem as one that requires a decision-making process. | 50 [50–75] | 50 [50–75] | 25 [25–25] |
|  | Clinicians used vague terms to talk about the index problems (cardiac arrest and/or respiratory failure): “if a disaster happens”, “if an undesirable event happens”, “if you are not doing well”.<br>Clinicians used vague and imprecise terms to talk about invasive mechanical ventilation: “put you on a machine”, “plug you to a fan”, “connect you to a machine”, “connect you to an artificial lung”. |  |  |
| 2. The clinician states that there is more than one way to deal with the identified problem (‘equipoise’). | 25 [25–50] | 25 [0–25] | 0 [0–25] |
|  | Only one intensivist explained the pros and cons of not attempting CPR or invasive mechanical ventilation.<br>Most discussions focused on the option to attempt an intervention: “If your heart is causing you trouble, the only option we have is to give it shocks and injections to try to restart it.”<br>The option not to attempt CPR was only presented once by an intensivist: “When it happens, we have two choices: we have the choice to do nothing and say: well the heart has stopped, life is over. And sometimes the choice we have is to try to restart the heart.”<br>The availability of palliative care in case of respiratory failure was never mentioned. |  |  |
| 3. The clinician assesses the patient’s preferred approach to receiving information to assist decision making (e.g., discussion, reading printed | 0 [0–0] | 0 [0–0] | 0 [0–0] |
|  | No intensivist assessed patients’ preferred approach to receiving information to assist decision making. |  |  |
| material, assessing graphical data, using videos or other media). |  |  |  |
| 4. The clinician lists ‘options’, which can include the choice of ‘no action’. | 25 [25–25] | 25 [0–25] | 25 [25–25] |
|  | See Item 2 |  |  |
| 5. The clinician explains the pros and cons of options to the patient (taking ‘no action’ is an option). | 25 [0–50] | 25 [25–25] | 25 [0–25] |
|  | See Item 2 |  |  |
| 6. The clinician explores the patient’s expectations (or ideas) about how the problem(s) are to be managed. | 25 [25–50] | 25 [25–50] | 50 [0–50] |
|  | In most cases, intensivists used euphemisms to talk about death and dying: “*to let you go*”, “*close the books*”, “*Jesus is coming to you*”.<br>Intensivists broadly explored patient’s fears and concerns about the consequences of CPR and invasive mechanical ventilation, but they did not explore their fears and concerns about the consequences of not attempting the interventions. |  |  |
| 7. The clinician explores the patient’s concerns (fears) about how problem(s) are to be managed. | 25 [25–75] | 25 [0–50] | 25 [25–50] |
|  | See Item 6 |  |  |
| 8. The clinician checks that the patient has understood the information. | 25 [25–25] | 25 [25–50] | 25 [25–25] |
|  | Most clinicians made minimal efforts with very simple verification terms to check if the information is understood: “*Ok?*”, “*Do you understand?*” |  |  |
| 9. The clinician offers the patient explicit opportunities to ask questions during the decision-making process. | 25 [25–25] | 50 [25–50] | 50 [25–50] |
|  | Clinicians used appropriate speech rhythm and asked if the patient had questions in general. However, they omitted to ask if patients had specific questions about the options discussed. |  |  |
| 10. The clinician elicits the patient’s preferred level of involvement in decision-making. | 0 [0–0] | 0 [0–0] | 0 [0–0] |
|  | No intensivist assessed the patient’s preferred approach to assist decision-making. |  |  |
| 11. The clinician indicates the need for a decision making (or deferring) stage | 25 [25–50] | 25 [25–25] | 0 [0–25] |
|  | Clinicians led patients to think about the options without naming the options or presenting the pros and cons of the options. Moreover, they were directive in making final decisions: “*The contract is: if a disaster [happens] we will use all interventions to keep you alive and we will take the time it takes to determine how your brain will be after all that. OK? So that's the contract.*” |  |  |
| 12. The clinician indicates the | 25 [0–50] | 0 [0–25] | 0 [0–75] |
| need to review the decision (or deferment). | Intensivists rarely indicated that the decision could be reviewed later while the patient was still capable. They often suggested delegating the decisions to the patient's surrogate decision maker if respiratory failure or cardiac arrest would occur. They often concluded discussions with patients that if cardiopulmonary resuscitation or if prolonged mechanical ventilation was needed, decisions about stopping life-sustaining interventions would be made with their surrogate decision makers if ever outcomes do not meet their overall quality of life expectations: *"Of course in your case, it would be very reasonable to say that we give you the maximum of chances and we can see if the results correspond with what you would like."* |  |  |
| **Total scores** | **25 [21–29]** | **21 [15–25]** | **19 [17–29]** |

*IQR* interquartile range, *n* count, *OPTION* refers to the *OPTION* scale score, *CPR* cardiopulmonary resuscitation

**Table 4.** Counts of observed quality indicators of end-of-life communications and decision-making.

|  | **Phase 1**<br>**(n = 7)** | **Phase 2**<br>**(n = 7)** | **Phase 3**<br>**(n = 7)** | **Total**<br>**(n = 21)** |
| --- | --- | --- | --- | --- |
| **Item observed** | **n, (%)** | **n, (%)** | **n, (%)** | **n, (%)** |
| 1. Prior to the discussion, documentation of the outcomes of advance care planning conversations (including any prior expressed wishes, diaries, and power of attorney documents) is consulted in the patient's medical record. | 4 (80) | 5 (83) | 3 (43) | 12 (67) |
| 2. Before the discussion, a member of the health care team provided the patient and/or their family with information about goals of care to look at before conversations with the doctor. | 0 (0) | 0 (0) | 0 (0) | 0 (0) |
| 3. During the discussion, the intensivist talked to the patient about a poor prognosis or indicated in some way that the patient had a limited time left to live. | 0 (0) | 0 (0) | 0 (0) | 0 (0) |
| 4. During the discussion, the intensivist asked if the patient had prior discussions or | 3 (43) | 4 (57) | 6 (86) | 13 (62) |
| had written documents about the use of life-sustaining treatments. |  |  |  |  |
| 5. During the discussion, the intensivist used information about goals of care to support the decision. | 0 (0) | 0 (0) | 0 (0) | 0 (0) |
| 6. After the discussion, documentation of goals of care is present in the medical record. | 7 (100) | 6 (86) | 7 (100) | 20 (95) |
| 7. After the discussion, the goals of care present in the medical record are consistent with the patient's stated preferences. | 5 (71) * | 5 (71) * | 6 (86) ** | 16 (76) |

*n* count, \* Two patients had unclear goals of care documented in their chart, \*\* One patient had unclear goals of care in their chart

**Table 5.** Goals of care discussions clinical encounter characteristics.

| Clinical encounters |  |  |  |  |
| --- | --- | --- | --- | --- |
|  | Phase 1<br>(*n* = 7) | Phase 2<br>(*n* = 7) | Phase 3<br>(*n* = 7) | Total<br>(*n* = 21) |
| **Use of the decision aid, *n* (%)** | 0 (0) | 0 (0) | 0 (0) | 0 (0) |
| **Use of other support documentation, *n* (%)** | 0 (0) | 0 (0) | 0 (0) | 0 (0) |
| **Presence of family members, *n* (%)** | 1 (14) | 2 (28) | 2 (28) | 5 (24) |
| **Genders of the dyads, *n* (%)** |  |  |  |  |
| Male patient/male intensivist | 1 (14) | 4 (58) | 4 (58) | 9 (43) |
| Male patient/female intensivist | 2 (28) | 2 (28) | 2 (28) | 6 (29) |
| Female patient/female intensivist | 3 (42) | 0 (0) | 0 (0) | 3 (14) |
| Female patient/male intensivist | 1 (14) | 1 (14) | 1 (14) | 3 (14) |
| **Encounters per intensivist: minimum-maximum** | 0–4 | 1–2 | 0–3 | 2–9* |
| **Duration (minutes), median (IQR)** | 12 (9–22) | 8 (6–14) | 17 (6–30) | 12 (7–18) |
| **Total number of words pronounced by patient and intensivist, median (IQR)** | 1600<br>(1114–2631) | 1882 (1373–1978) | 2131 (1006–4127) | 1882<br>(1064–2721) |
| **Percentage (%) of words pronounced by the intensivist,** | 57 (53–72) | 68 (60–82) | 66 (67–89) | 66 (57–85) |
| **median (IQR)** |  |  |  |  |

*IQR* interquartile range, *n* count, \* One intensivist conducted 9 of the 21 encounters (43%)

### 3.3 Quality of end-of-life communication and decision-making

Quality of end-of-life communication and decision-making was similar across all phases (Table 4). Intensivists performed well on Item #6 (*GCD documentation in the medical record*), but poorly on items #2 (*Information about goals of care provided before GCD*), #3 (Poor prognosis shared with patients) and #5 (*Information about goals of care provided to support the decision during GCD*) across all phases of study.

### 3.4 Goals of care discussions characteristics

GCD characteristics are described in Table 5. None of the intensivists used the DA during the discussions included in this study.

### 3.5 Elderly patients’ most frequent misunderstandings

Patients confused the concept of goals of care with the completion of legal documents (e.g., power of attorney, will). After the intensivist introduced the need to discuss their goals of care, patients replied that they had already discussed these with their notary:

> *“At the notary’s office we both wrote that we did not want futile aggressive care”* [translated from French*: acharnement thérapeutique*]

The term “*acharnement thérapeutique*”, translated in English as “*futile medical care*”, is used both in plain language and on legal documents. It does not have a precise medical definition meaning that clinicians still need to clarify their patient’s wishes and define what futile care means for each individual.

A few patients (*n* = 4) who stated that they decided with their notary not to receive futile aggressive end-of-life care decided with their intensivist to change their previous decision and now wanted to prolong their life with all necessary means.

Three patients also confused GCD with discussions about organ donation:

> *“Yes, it’s for organ donation. It’s signed. It’s my wife who does that kind of stuff.”*

Another common misunderstanding we identified was that patients under evaluated or did not understand the possible consequences of CPR. While some patients (*n* = 3) wished to receive all necessary interventions to prolong their life (including CPR) they also refused to be “*connected to a machine*” which demonstrates patients’ misconception about CPR that usually includes being mechanically ventilated.

### 3.6 Elderly patients’ most frequent concerns

Seven patients feared losing functional autonomy and having to move to a long-term care facility following life-sustaining treatments:

> *“I do not want to be paralyzed and have to go to a long-term care facility. I would rather die than go to a long-term care facility.”*

Another concern (*n* = 7) was losing cognitive capacities and surviving in a vegetative state:

> *“I do not want to be a vegetable. This is the worst thing that can happen to someone.”*

Another patient expressed the desire for medically-assisted death if ever he became severely incapacitated and incapable of making his own decisions:

> *“I’m not interested, if I end up too messed up and need to go to a long-term care facility. If the law exists, I want euthanasia.”*

Some patients (*n* = 6) requested to receive all possible interventions to prolong their life in case of a life-threatening emergency, but also expressed wanting the medical team to let them die if they were to survive in a vegetative state. Consequently, a common way of concluding these discussions by intensivists was to offer CPR and mechanical ventilation and postpone any difficult decisions until after the interventions had occurred and then to delegate the decision to a surrogate.

> *“So, we will do everything we can to keep you alive, but if we see that you are going to end up in a state that you would not wish for, we will stop.”*

This frequent decision-making postponement strategy translated into written physician orders that mandated life-sustaining therapy as long as a return to previous life was possible.

### 3.7 Evaluation of the training, barriers and facilitators to the use of shared decision making and decision aid

All intensivists completed the online training and participated in the debriefing sessions after Phase 2 and after the study’s completion. Participants perceived that the overall quality of the online training, its organization, clarity of its content, clarity of its instructions about using the decision aid was excellent (Appendix 6). Intensivists’ sense of self-efficacy in SDM and use of the DA increased after the training (Appendix 6: Table A3). During the post-study debriefing session, many barriers and facilitators to SDM and the DA were identified by intensivists (Appendix 7). The main barriers were: (1) physician’s attitude (e.g., difficulty in changing habits, fear to generate patient anxiety, difficulty in talking to patients about death), (2) challenges for optimal care in the ICU (e.g., lack of time, and difficult, complex and urgent clinical situations), (3) misunderstanding of clinicians about SDM and DAs, (4) lack of practical training, and (5) change management issues. The main facilitators were: (1) engaged physicians’ inclination to use a patient-centered approach, (2) clearer and more accessible DA, (3) earlier, more frequent and better public and patient education about GCD before entering the ICU, ideally in the primary care setting, (4) training of all health professionals and developing an interprofessional approach to GCD in the ICU, (5) access to evidence-based tools, (6) family and surrogate decision-maker inclusion in the discussions, (7) support from all health professionals in the ICU, and (8) patients’ positive and receptive attitude to having GCD.

## 4 Discussion

Our main objective was to explore the impact of a context-adapted DA and a SDM training on intensivists’ involvement of elderly patients in SDM about their goals of care and the quality of GCD. Our exploration generated four main findings. First, even though all the eligible intensivists completed the online SDM training and felt more empowered to lead SDM and use the DA after the online training, none of the intensivists used the DA with their patients. Second, our small sample size prevented us from making conclusions about the impact of the online SDM training on the involvement of patients in SDM or on the quality of GCD. However, we did identify barriers that may prevent intensivists to lead high-quality GCD and to better involve patients in SDM. Third, the Conceptual Framework for Improving End-of-life Communication and Decision-making allowed us to evaluate quality indicators relevant to critical care and pose areas for quality improvement. Fourth, we provide new evidence about the use of recording real-world, sensitive discussions of goals of care and end-of-life questions in the ICU.

We add to the existing literature about the challenges in studying SDM and GCD in the ICU. First, many patients admitted to the ICU are incapable of engaging in SDM because of their medical condition (e.g., experiencing delirium or have been intubated). As such, we underline the importance of earlier and better GCD before being admitted to the ICU [36]. Although our ICU is a closed unit under the leadership of a dedicated intensivist, reticence also remained among participating intensivists to engage in a new GCD with patients if a discussion had already taken place with a different admitting or consulting physician [37–39]. This also aligns with existing research highlighting how physician rotation and shift changes in the ICU complicate involving the primary physician in GCD discussions [40]. Encouraging all members of the care team involved in the discussion—irrespective of their status as attending, ordering or consulting physicians—to discuss the goals and preferences of their patient may ensure GCD can occur even if the primary physician is not present.

Team communication fosters a deeper understanding of changing patient care goals, especially at the end of life. When team members may comfortably and candidly share their ideas, it ultimately benefits patient care. Refinement of the DA and its training to explicitly incorporate a team-based approach, particularly an interprofessional team-based approach may scaffold ideal participation of the care team. This aligns with a promising trend in end-of-life care, where teams facilitate GCDs and ensure that care aligns with patient preferences [41–43]. Intensivists also identified the need to incorporate more health professionals in the decision-making process which can improve care and facilitate the SDM process [44].

Second, although intensivists highly value the importance of leading high-quality GCD, they still avoided talking about death and did not use a DA that presented death as a potential outcome.

Only one intensivist discussed the option not to attempt resuscitation and none presented the availability of a palliative care approach led by the intensive care team. The potential reasons for this were identified in a previous study about limits to end-of-life communications in Canadian hospitals: uncertainty about what counts as end-of-life, active avoidance of end-of-life conversations, feeling that everything should be done until nothing can be done [21]. In our study, intensivists also highlighted the fear of creating more anxiety for both them and for their patients in using our DA [45]. These barriers and others such as confusion about which professional should initiate the conversation, lack of organizational support, lack of palliative care skills for intensivists are currently being addressed by interventions such as the Serious Illness Conversation Guide centered around better communication and best practices in palliative care [46–48].

Third, our analysis identifies the use of euphemisms and metaphors for death and dying, which are an indirect way of communicating the topic and occurs in other medical specialties that deal with end-of-life care [49]. For example, an oncology study found that only 52% of oncologists explicitly discussed death with patients with advanced forms of cancer [50]. Although empathy must remain central in GCD [51], using implicit language can prevent patients from taking appropriate actions and correctly weigh the risks and benefits of the available options [51]. In our study, it is unclear whether our participants understood that refusing CPR/mechanical ventilation could lead to death because the palliative care options were not stated, nor discussed. These represent potential areas for improving intensivists SDM skills about goals of care and end-of-life and implementation strategies to explore in future projects aiming at improving GCD in the ICU.

Fourth, many misunderstandings and concerns persist among patients about the concept of goals of care, the invasive interventions available in the ICU and their associated risks and benefits.

Patients contribute to delaying decision-making as many prefer to defer difficult decisions to their surrogate decision makers and/or their medical team once they become unable to make decisions [5,8]. In our study, six patients expressed the wish to receive all possible life-sustaining interventions, but also expressed the desire to die without suffering if they were to survive with severe deficits. This puts the burden on surrogate decision makers who need even more guidance to make these decisions because they do not always know what their loved ones want and as such, many conflicts arise. Despite this risk of conflict, intensivists in our study recognize the importance of involving surrogate decision makers earlier in GCD to address this problem.

Fifth, our study was underpowered to demonstrate that training in SDM and the use of a DA could increase patient involvement or the quality of GCD. While physicians expressed satisfaction with the training and the tool itself, it became clear that knowledge and training alone is insufficient to drive changes in clinical practice. This aligns with a previous systematic review which identified various barriers beyond knowledge, including system limitations and physician-specific challenges [52]. Our own physician feedback mirrored these findings, highlighting time constraints, barriers regarding attitudes and behaviors, and the fast-paced nature of critical care environments as significant hurdles [53]. The ICU setting presents unique challenges, with physicians often feeling a sense of sole responsibility and pressure to navigate emotionally charged discussions with patients and families [54]. To improve the implementation of SDM in the ICU, a shift beyond simply providing discussion tools is necessary. Instead, future efforts should focus on fostering an environment that supports physicians, allows for adequate time for patient interactions, and demonstrates the positive impact of their engagement with these tools [55]. Similarly, creating conditions that promote patient comfort, openness, and understanding during discussions is crucial for achieving meaningful SDM in critical care.

Moreover, the OPTION scale allowed us to highlight areas of potential improvement for intensivists’ SDM communication skills about goals of care and end of life. Although our median OPTION scores were low, they are similar to the baseline OPTION scores reported in a systematic review assessing interventions to improve the involvement of patients in SDM (the OPTION score mean was 23% (SD = 14%) [30]. We found that intensivists did not ask patients whether they wanted to play an active or passive role in decision making: an essential element of SDM that was covered in our training material. This highlights a need for continued training and quality improvement about SDM in the ICU [29,31].

Sixth, the Conceptual Framework for Improving End-of-life Communication and Decision-making allowed us to evaluate quality indicators that are essential to critical care. While the OPTION scale is a general tool used to evaluate all kinds of decisions, this framework measures end-of-life communication and decision-making specifically. We struggled with measuring the concept of *consistency* used in the framework [7]. In total, 76% of the goals of care present in the medical records after the discussion were consistent with the patient’s stated preferences.

However, patients’ preferences are not always consistent throughout GCD and often conflict with each other. For instance, some patients wanted every intervention to prolong their life but did not want to be connected to a machine. This highlights the fact that intensivists may not be not clear enough during their GCD. Other intensivists are more direct in presenting goals of care that they believe are most appropriate for their patient without presenting all the available options based on their clinical judgment and their understanding of their patient’s prognosis and preferences. This potentially increases apparent consistency in the medical chart, but makes it harder to determine if the patient understood that some options were not available and if decision-making was consistent with the patient’s preferences. More research is justified to determine how and when to measure the consistency of physician orders for life-sustaining therapies.

Finally, our study sheds new light on the use of videorecording sensitive goals of care and end-of-life questions in the ICU. Only one participant preferred audio recording over video recording. None of the intensivists refused to be video recorded. The acceptance rate in other studies was also high, confirming that video and/or audio recording can be a valuable tool to study decision making and identify quality improvement opportunities [51,56–62]. Here, videorecording offered additional opportunities to better understand physician-patient interactions and better identify patient’s emotional reactions and misunderstandings to different physician statements. Further studies are needed to explore the use of videorecording to conduct other in-depth qualitative analyses of patient misunderstandings and help improve physician communication skills.

### 4.1 Limitations

Our study has several limitations. First, our main hypothesis was that our intervention (a context-adapted DA combined with training clinicians about SDM) would increase clinicians’ adoption of SDM and the quality of GCD. Though all the intensivists completed the online training, no intensivists used the DA during the clinical encounters we captured. Our results may have been different if the DA had actually been used with patients. This DA was tested in the same ICU that it had been developed in. Even with user centered design and active engagement of local users, our DA was still not used. Future work will have to address barriers we identified in implementing our DA to improve our online training module. Second, our study was underpowered with only five participating intensivists in a single intensive care unit. This prevented us from performing statistical testing and modeling to adjust for time-dependent correlations between individual intensivists in each phase. Third, we conducted our study in a pragmatic, real-world environment with a limited amount of time and availability to capture the encounters (2 months per phase). As such, we could not control the number of encounters conducted per intensivist, per phase allowing for over-representation of certain intensivists throughout the 3 phases. Finally, we were not able to capture discussions occurring off-hours.

## 5 Conclusion

The adoption of SDM was low and many areas remain to improve the quality of GCD in the ICU. A larger sample size is needed to better evaluate the impact of a context-adapted DA and SDM training and on the adoption of SDM by intensivists about GCD. Our qualitative results highlight many patient misunderstandings that intensivists can address and areas where they can improve their SDM communication skills. More qualitative research is needed to understand the role of DAs to support SDM and better involve older adults in goals of care decisions within the ICU.

## Supporting information

English Supplement (Appendices 1 to 9)

## Data Availability

All anonymized data produced in the present study are available upon reasonable request to the authors.

## List of abbreviations

DA: decision aid
SDM: shared decision-making
GCD: goals of care discussions
ICU: intensive care unit
ACCEPT: Audit of Communication, Care Planning, and Documentation
IQR: interquartile range
CPR: Cardiopulmonary resuscitation
StaRI: Standards for Reporting Implementation Studies
COREQ: Consolidated criteria for reporting qualitative research

## Acknowledgements

We express our gratitude to Annie Leblanc, William Witteman, Maude Linteau, Louise Sauvé, Sarah Alameddine, Daniel Paré, and Pascal Smith, for their contributions and support in enhancing the quality of this project. Simultaneously, we pay a heartfelt tribute to our patient-partner Paulette Archambault (1928-2020) and our colleague Jennifer Kryworuchko (1968- 2019), whose invaluable contributions will forever resonate in this work. The online training was developed by Ariane Plaisance, Frédérick Noiseux, Annie LeBlanc, France Légaré, Patrick Plante, Louise Sauvé, Jennifer Kryworuchko, Diane Tapp, Emmanuelle Bélanger, Mark H Ebell, Todd Gorman, Julien Turgeon, Alexis Turgeon, Holly Witteman, Hubert Marcoux, Anne-Marie Boire-Lavigne, Daniel Paré, Sarah Alameddine, Tom van de Belt, Christian Chabot, Paulette Bergeron, Marie-Josée Poiré, Carrie Anna McGinn, Audrey Dupuis, Hugues Vaillancourt, Rebecca Francois, and Patrick M Archambault.

## Declarations

### Ethics approval

The project was approved by the Centre intégré de santé et de services sociaux de Chaudière-Appalaches (CISSS-CA) Research Ethics Board (Certificate #2017-001).

### Consent to participate

A member of the research team obtained written consent to participate from each patient. Family members wishing to participate also completed a consent form. Patients, family members, and physicians were made aware that it would not be possible to identify individual participants in the published results.

### Consent for publication

Not applicable.

### Availability of data and material

The datasets used and/or analyzed during the current study are available from the corresponding author on reasonable request.

### Code availability

The code generated and used in the analysis of the study datasets is available from the corresponding author on reasonable request.

### Authors’ contributions

All authors interpreted the results, wrote the final manuscript, and agreed to its published version.

