## Supplementary material for "Exploring the impact of a context-adapted decision aid and online training about shared decision making about goals of care with elderly patients in the intensive care unit: a mixed-methods study": English Supplement (Appendices 1 to 9)

**Emmanuelle Bélanger**, PhD^[13]^, **Mark H Ebell**, MD, MS^[14]^, **Christian Chabot**, Patient Partner^3^,

**Tom van de Belt**, PhD^[15]^, **Alexis F Turgeon**, MD, MSc, FRCPC^[16],^^[17]^, **Patrick M Archambault**, MD, MSc, FRCPC^3,4,5,6^

**Corresponding author:**

**Patrick M Archambault**

Department of Family Medicine and Emergency Medicine, Université Laval.

Vandry Pavilion, 2325, rue de l'Université, Québec, QC, Canada, G1V 0A6.

1. Sciences de la santé, Université du Québec à Rimouski (UQAR), Lévis, Québec, Canada
2. Institut universitaire de cardiologie et de pneumologie de Québec (IUCPQ), Department of anesthesia and critical care, Faculté de médecine, Université Laval, Québec, Canada
3. VITAM - Centre de recherche en santé durable, Québec, Québec, Canada
4. Centre de recherche en santé durable, Centre intégré universitaire de santé et de services sociaux de la Capitale-Nationale (CIUSSS-CN), Québec, Canada
5. Department of Family Medicine and Emergency Medicine, Faculté de médecine, Université Laval, Québec, Canada
6. Centre de recherche intégrée pour un système apprenant en santé et services sociaux, Centre intégré de santé et services sociaux de Chaudière-Appalaches (CISSS-CA), Lévis, Québec, Canada
7. Centre hospitalier universitaire de Québec-Université Laval (CHUL), Québec, Québec, Canada
8. Faculté de médecine, Université Laval, Québec, Québec, Canada
9. Université TÉLUQ, Québec, Québec, Canada
10. Centre de recherche et d’innovation en technologie éducative du Québec (i-TEQ)
11. Faculté des sciences infirmières, Université Laval, Québec, Québec, Canada
12. Centre de recherche hospitalier universitaire de Québec-Université Laval (CHUL; Oncology Axis), Québec, Québec, Canada
13. Center for Gerontology and Healthcare Research, Brown University School of Public Health, Providence, Rhode Island, USA
14. Department of Epidemiology and Biostatistics, College of Public Health, University of Georgia, Athens, Georgia, USA
15. Research Group Technology for Health, HAN University of Applied Sciences, Nijmegen, The Netherlands
16. Centre de recherche hospitalier universitaire de Québec-Université Laval (CHUL; Population Health and Optimal Practices Unit), Québec, Québec, Canada
17. Department of Anesthesiology and Critical Care Medicine, Division of Critical Care Medicine, Faculté de médecine, Université Laval, Québec, Canada

### Appendix 1. Training program content

The full content of this training program is available at https://fpdp.archambaultlab.ca/.

This online training program was conceived by a group of SDM and intensive care medicine experts. This module was designed to last approximately one hour.

The online training program was followed by an in-person debriefing session with the principal investigator and a research professional.

The online training program is centered on SDM in the ICU, the available decision aid tools and a conceptual “4 step encounter” framework to help clinicians apply the principles of SDM to goals of care discussions.

The training program contains 3 modules:

**Module 1 – Shared decision making**

This section presents the concept of SDM. It gives a perspective on the different roles the physician can have in decision making, from a paternalistic approach to an autonomist approach. It presents the 9 elements of SDM according to Makoul and Clayman [29].

**Module 2 – Decision aids**

This section presents three different decision aids available to help the clinician and patients during goals of care discussions.

- The first decision aid is a paper booklet providing information about CPR and mechanical ventilation, and providing questions for the patient to engage in a reflexive process concerning his/her wishes.
- The second decision support tool is the online GO-FAR calculator. This calculator helps predict the probability of a favorable/unfavorable neurological outcome after undergoing in-hospital cardiopulmonary resuscitation tailorable to each patient’s risk factors.
- The third decision aid is a video version of our paper decision aid that covers the same topics as the paper booklet.

**Module 3 – The four-step encounter**

This module proposes a four-step approach to any goals of care discussion.

Step 1- Engage. Present the goal of the discussion and its importance. Obtain the consent of the patient to pursue it.

Step 2- Empathy. Explore the patient’s point of view of his global health condition. Explore his level of autonomy and wishes.

Step 3- Educate. Agree on an appropriate level of care.

Step 4- Enlist. Summarize the decision, conclude the discussion.

The training program also featured a video presenting a simulated clinical encounter using the communication skills presented during the module.

Finally, the online module concludes with a post-test evaluation of knowledge acquired and a satisfaction questionnaire. Appendix 6 presents the results of the satisfaction questionnaire.

### Appendix 2. Feedback session verification list.

These slides were translated from the original French for an interactive discussion session held on November 6^th^, 2017. Please contact the corresponding author for the original materials in French.

**What is your understanding of the nine elements that make up shared decision-making?**

Do you have any questions/comments about this section?

2

**Defining the issue of discussions on levels of care in the current context**

Do you have any questions/comments about this section?

1

**How do you see using these tools in your everyday work?**

6

**Presentation of the GO-FAR calculator**

Do you have questions about this tool?

5

**Presentation of the paper context-adapted decision aid**

Do you have questions about this tool?

4

- Define/explain problem
- Present options
- Discuss benefits/risks/costs
- Clarify patient values/preferences
- Discuss patient ability/self-efficacy
- Discuss doctor knowledge/recommendations
- Check/clarify patient understanding
- Make or defer a decision
- Arrange follow-up

3

**Do you have any comments on the visual or practical aspects of the online training session?**

8

**According to the four steps for conducting a goals of care discussion with the patient, do you have any questions or comments to share?**

7

### Appendix 3. OPTION scale in English.

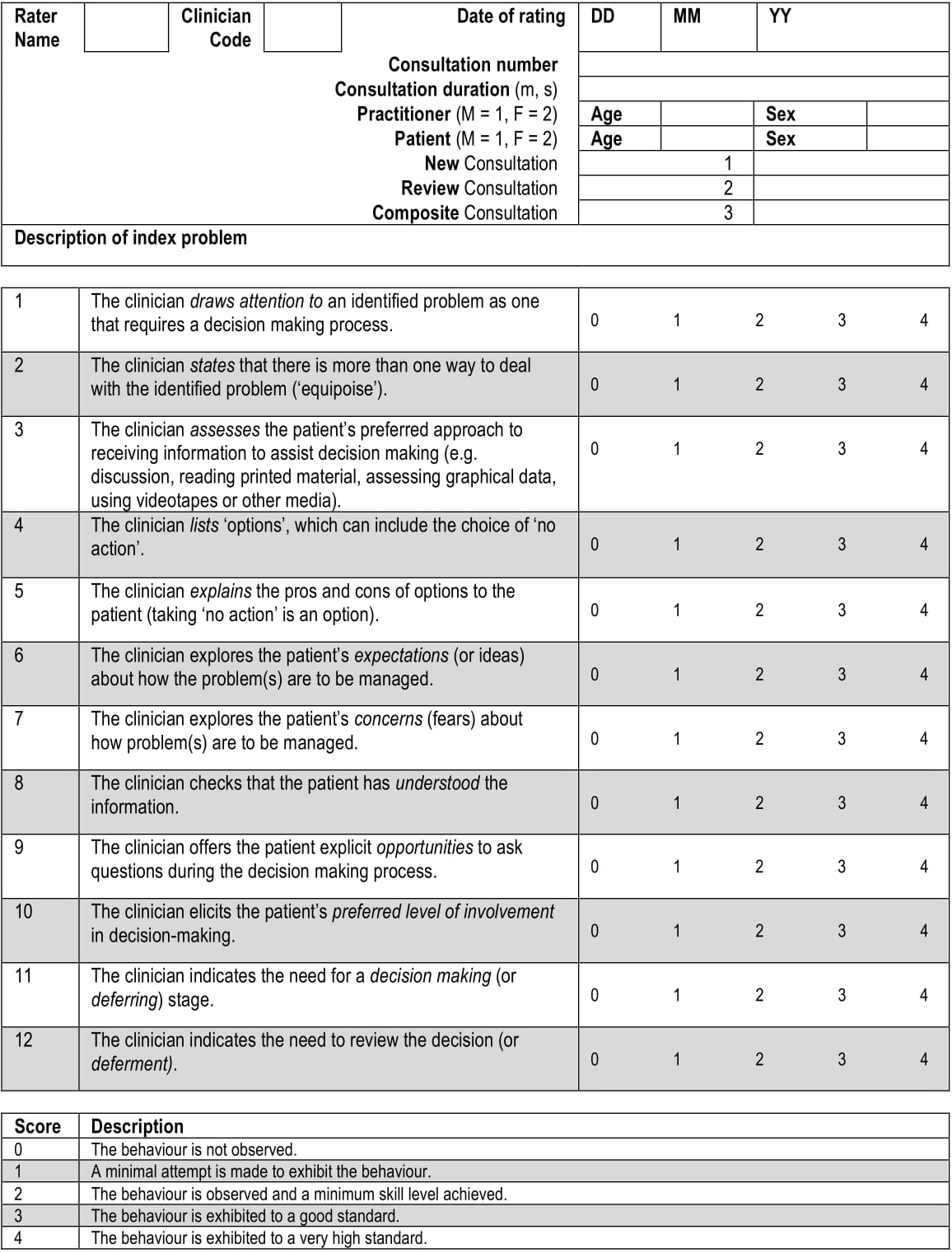

Please contact the corresponding author for the French version of this scale used in this study. You may also consult this [Critical Synthesis Package](https://www.mededportal.org/doi/full/10.15766/mep_2374-8265.10128) for more information on using the scale, and for more instrument formats.

### Appendix 4. Four-item OPTION scale coding guide based on the health issues that were included in the decision aid

**Item 1. The clinician draws attention to an identified problem as one that requires a decision-making process.**

0 = No attempt is made to draw attention to the need to engage in a decision process. (There is no clearly identified problem, or at least no clear indication that a decision has to be made.)

*Indice: The clinician doesn't mention any of the index interventions (CPR and mechanical ventilation), and doesn’t mention the need for a decision to be made.*

1 = Very brief or superficial attempts to draw attention to the need for a decision process to be engaged.

*Indice: The clinician identifies one or both of the index interventions (CPR and/or mechanical ventilation), but doesn’t clearly express that a decision has to be made*

2 = Basic level of competence: the clinician draws the attention to a problem that requires a decision process.

*Indice: The clinician identifies one or both of the index interventions (CPR and/or mechanical ventilation), and clearly expresses that a decision has to be made*

3 = The clinician highlights the need for a decision process to be engaged.

*Indice: The clinician clearly presents both the interventions and clearly expresses that a decision has to be made about those interventions.*

4 = The competence is demonstrated at a high level. (Additional explanations, the clinician incites the patient to recognize the importance of engaging himself in the decision process)

*Indice: The clinician presents both the intervention and gives supplementary explanations on the importance of engaging the patient in the decision process.*

**Item 2. The clinician states that there is more than one way to deal with the identified problem (‘equipoise’).**

0 = The clinician does not mention that there is more than one way to deal with the problem.

*Indice: The clinician does not mention that there is more than one option.*

1 = Superficial attempts to communicate the existence of more than one option.

*Indice: The clinician mentions that there is more than one option for one of the index interventions (CPR or mechanical ventilation). He can use words like “things”, “ways”, “manners” or any synonym expressing the idea that there is more than one way to deal with the problem.*

2 = Basic level of competence: the clinician states that all options are valid and need to be considered in a detailed manner.

*Indice: The clinician exposes all the available options for both the index interventions (CPR and mechanical ventilation). The clinician states that all options are equal and that they need to be considered in a detailed manner.*

3 = The clinician explains the concept of “equipoise”, and that all options have advantages and disadvantages that need to be considered.

*Indice: The clinician defines the concept of “equipoise” or specifies that all options have upsides and downsides.*

*Indice:* The clinician explains why more than one option exists for the interventions. He explains why every option is available.

*Indice: The clinician explains why more than one option exists and why every option is available.*

**Item 3. The clinician assesses the patient’s preferred approach to receiving information to assist decision making (e.g., discussion, reading printed material, assessing graphical data, using videotapes or other media).**

0 = The behavior is not observed.

*Indice: The clinician does not mention that more than one way to receive the information exists.*

1 = A minimal attempt is made to exhibit the behavior.

*Indice: The clinician mentions that there is more than one way to receive the information without mentioning such ways, or gives the patient the decision-making aid without asking his preference.*

2 = Basic level of competence: the clinician asks the patient in which way he prefers to receive the information.

*Indice:The clinician mentions that there is more than one way to receive the information and asks what is the patient’s preference.*

3 = The behavior is exhibited to a good standard. The clinician states that there is more than one way to receive the information and gives documentation to consult outside of the encounter.

*Indice: The clinician presents all the available ways to receive the information and, according to the patient’s preference, gives the decision aid. He does not explore the document with the patient.*

4 = The behavior is exhibited to a very high standard. The clinician gives many examples of available ways to receive the information, and thereafter gives the patient the opportunity to choose his preferred medium (paper, phone screen, etc.).

*Indice: The clinician exposes all the available media, and gives the patient the choice of his preferred manner to receive the information.*

**Item 4. The clinician lists ‘options’, which can include the choice of ‘no action’.**

0 = The behavior is not observed.

*Indice: The clinician doesn’t present any options.*

1 = A minimal or superficial attempt is made to list the available options.

*Indice: The options are presented, but there is no list or structure, and the option of “not doing anything” is not offered*.

2 = Basic level of competence: The clinician lists all the available options, using words like “or”, “instead” to clearly distinguish them.

*Indice: There is a clear list of the available options and they are clearly distinct one from another.*

3 = The clinician carefully lists all the available options, including the option of “not doing anything” or reporting the decision.

*Indice: There is a clear list of the available options and they are clearly distinct from one another, and the option of “not doing anything” or reporting the decision is offered.*

4 = The behavior is exhibited to a very high standard.

*Indice: The clinician presents all the available options in a clear and detailed manner. He details the situations that require CPR and mechanical ventilation and gives details about the duration of CPR and mechanical ventilation if needed.*

**Item 5. The clinician explains the pros and cons of options to the patient (taking ‘no action’ is an option).**

0 = No explanations are given.

*Indice: The clinician doesn’t present any of the pros or cons.*

1 = The clinician doesn’t give explanations on more than one option.

*Indice: The clinician presents information about one option concerning one or both of the index interventions (CPR and/or mechanical ventilation). For example, the clinician presents the pros and cons of mechanical ventilation but doesn’t mention the pros and cons of refusing this intervention.*

2 = Basic level of competence: the clinician provides details about the pros and cons of the options.

*Indice: The clinician presents information about at least two options concerning one or both of the index interventions (CPR and/or mechanical ventilation). For example, the clinician presents the pros and cons of mechanical ventilation and the pros and cons of refusing it.*

3= The behavior is exhibited to a good standard.

*Indice: The clinician goes further in the explanations. Exhaustive presentation or in the form of an organized list of the pros and cons concerning one or both the index interventions (CPR and/or mechanical ventilation). He mentions that refusing one of the index interventions involves death.*

4 = The behavior is exhibited to a very high standard.

*Indice: The clinician presents all the pros and cons of both interventions (CPR and mechanical ventilation). He mentions that refusing one of the index interventions involves death.*

**Item 6. The clinician explores the patient’s expectations (or ideas) about how the problem(s) are to be managed.**

0 = No attempt is made to explore the patient’s expectations.

*Indice:* The clinician doesn’t attempt to verify the patient’s ideas.

1 = Superficial attempt to explore the ideas or expectations of the patient on resolving the problem.

*Indice:* The clinician presents some examples that lead the patient to reflection. He opens the door to the reflection on the expectations about the management of the problem.

2 = Basic level of competence: The clinician explicitly asks the patient about his expectations concerning the solving of the problem. He explores the ideas and expectations using open questions, bringing up things in common, and by using pauses.

*Indice:* The clinician explores the expectations. He explicitly asks the patient about his expectations concerning the problem.

3 = The behavior is demonstrated and leads to supplementary questions to clarify the patient’s ideas and expectations. The behavior is demonstrated at a good level.

*Indice:* The clinician explores in a deeper manner the expectations and asks supplementary questions.

4 = The behavior is exhibited to a very high standard and the patient’s perspective is explored.

*Indice:* The patient’s perspective is thoroughly discussed.

**Item 7. The clinician explores the patient’s concerns (fears) about how problem(s) are to be managed.**

0 = No attempt is made to explore the patient’s concerns.

*Indice:* The clinician doesn’t explore the patient’s concerns.

1 = Superficial attempts to identify the patient’s concerns about the management of the problem.

*Indice:* The clinician invites reflection by his examples. He prompts the patient to think about his or her concerns. He reformulates the concerns that he seems to perceive in the patient’s speech instead of directly questioning him.

2 = Basic level of competence. The clinician explicitly asks the patient about his or her concerns concerning the interventions potentially needed to solve the problem. He explores concerns using open questions, common ways, and pauses.

*Indice:* The clinician explicitly asks the patient about his or her concerns.

3 = The behavior is exhibited and leads to supplementary questions to clarify the concerns.

*Indice:* The clinician provides many examples, explanations. He asks supplementary questions after asking the patient explicitly about his or her concerns.

4 = The behavior is exhibited to a very high standard: the patient’s concerns are discussed and considered.

*Indice:* In addition to exploring the patient’s concerns, the clinician adjusts the options according to them, in a goal of tailoring the possible interventions to them.

**Item 8. The clinician checks that the patient has understood the information.**

0 = No attempt is made to check that the patient has understood the information.

*Indice:* The clinician doesn’t check if the patient has understood the information.

1 = Superficial attempt to check that the patient has understood the information.

*Indice:* The clinician makes a minimal effort to check if the information is understood. Superficial questions like “Alright?”, “Ok?” are asked at least once during the discussion.

2 = Basic level of competence. The clinician explicitly asks the patient if he has understood the information provided.

*Indice:* The clinician explicitly asks if the patient has understood, with questions like “Is it clear?”, “Do you understand?”.

3 = The clinician checks the patient’s comprehension using affirmations like “I would like to make sure that you have understood the information concerning the available options. Can you tell me what you understood about…?”.

*Indice:* The clinician asks the patient to reformulate in his words.

4 = The behavior is exhibited to a very high standard.

*Indice:* The clinician asks the patient to reformulate in his words and ascertains throughout the discussion that the information given is clearly understood and makes sure he and the patient share the same comprehension.

**Item 9. The clinician offers the patient explicit opportunities to ask questions during the decision-making process.**

0 = No attempt to give the patient opportunities to ask questions is made.

*Indice:* No space is left to the patient to ask questions (no pauses, quick speech rhythm, interrupts the patient).

1 = The clinician uses pauses, or gives the patient the opportunity to ask questions by using an appropriate speech rhythm.

*Indice:* The clinician doesn’t interrupt the patient, uses a minimum of pauses, he uses a speech rhythm that demonstrates a minimum of welcoming to questions.

2= Basic level of competence. The clinician explicitly gives the patient the opportunity to ask his/her questions.

*Indice:* The clinician uses pauses and an appropriate speech rhythm, and asks an explicit question (“Do you have questions?”).

3 = The clinician is more explicit and asks the patient if he/she has questions about the options and the solution to the problem discussed.

*Indice:* The clinician explicitly asks the patient if he/she has questions concerning the concrete information given.

4 = The behavior is exhibited to a very high standard. The clinician gives the patient time to answer and checks if the patient has additional questions.

*Indice:* The clinician explicitly asks the patient if he/she has questions concerning the concrete information given, leaves time for the patient to formulate his/her interrogations and makes sure he/she has answered all the questions.

**Item 10. The clinician elicits the patient’s preferred level of involvement in decision-making.**

0 = No attempt is made to elicit the patient’s preferred level of involvement.

*Indice:* The clinician doesn’t ask the patient which role he/she wants to have in the decision-making process.

1 = Superficial attempt to elicit the patient’s preferred level of involvement in the decision-making process.

*Indice:* The clinician superficially implies that the patient can chose the role he/she wants to have in the decision-making process.

2 = Basic level of competence. The clinician explicitly asks the patient which role he/she prefers to have in the decision-making process.

*Indice:* The clinician explicitly asks the patient which role he/she prefers to have in the decision-making process, without explaining the different roles. He mentions the possible roles the patient can have without explaining them.

3 = The clinician offers additional explanations and evaluates what is the patient’s preferred role.

*Indice:* The clinician explains the different roles the patient can have in the decision-making process and precises the levels of responsibility the patient can have.

4 = The clinician asks the patient which role he/she prefers to have in the decision-making process in an understandable way. The clinician shows that he/she is aware of the decisional responsibility that is expected from the patient.

*Indice:* The clinician explicitly asks the patient which role he/she prefers to have in the decision-making process, in a way that makes the patient understand that it is important for him that he can choose his level of involvement in the decision.

**Item 11. The clinician indicates the need for a decision making (or deferring) stage.**

0 = The clinician doesn’t clearly indicate that the time has come to take a decision (or defer it).

*Indice:* The clinician doesn’t say anything concerning the fact that a decision has to be made (or deferred).

1 = Superficial attempt to indicate the need for a decision to be made.

*Indice:* The clinician leads the patient to think about the decision without clearly stating that a decision has to be made. For example, the clinician can say “Today, we are going to talk about what could happen if your heart stopped beating”.

2 = Basic level of competence. The clinician clearly states the need for a decision to be made. He can use statements like “Maybe the time has come to make a decision about what we should do”.

*Indice:* The clinician states that the time has come to make a decision. He uses the word “decision” or “choice”.

3 = The behavior is exhibited to a good standard.

*Indice:* The clinician clearly expresses that after all the information is given, it is time to make a decision or defer it if the patient wants to think about it or discuss it with relatives.

4 = The clinician allows for a transition from the considering of the pros and cons, to the patient’s ideas, preferences and fears, to the final decision.

*Indice:* The clinician resumes all the options. He allows for the patient to express his/her concerns and ideas and then signals that it is time to make a decision (or defer it).

**Item 12. The clinician indicates the need to review the decision (or deferment).**

0 = No attempt is made to indicate the need to review or defer the decision.

*Indice:* The clinician doesn’t mention that the patient can review his/her decision.

1 = Superficial attempt to indicate the need to review or defer the decision.

*Indice:* The clinician implicitly states that the patient can reconsider his/her decision. He mentions the possibility of “meeting again” without mentioning the decision.

2 = Basic level of competence. The clinician clearly indicates that the patient should be met again to review the decision.

*Indice:* The clinician indicates that the decision can be reviewed if wished (he/she offers the possibility).

3 = The behavior is exhibited to a good standard.

*Indice:* The clinician mentions that the patient can reconsider his/her decision and discuss it again with him/her or a member of the team if desired.

4 = The behavior is exhibited to a very high standard. It is made very explicit that the decision can be reviewed.

*Indice:* The clinician strongly encourages the patient to think about his/her decision and clearly mentions that it can be reconsidered and rediscussed.

### Appendix 5. Audit of Communication, Care Planning, and Documentation (ACCEPT) quality indicators used in our study

1) “*before the discussion, a member of the healthcare team provided the patient and/or their family with information about goals of care to look at before conversations with the doctor*”

2) “*during the discussion, the intensivist talked to the patient about a poor prognosis or indicated in some way that the patient has a limited time left to live*”

3) “*during the discussion, the intensivist asked if the patient had prior discussions or has written documents about the use of life-sustaining treatments*”

4) “*during the discussion, the intensivist used information about goals of care to support the decision*”

5) “*prior to the discussion, documentation of the outcomes of advanced care planning conversations (including any prior expressed wishes, diaries, and power of attorney documents) was consulted in the patient’s medical record*”

6) “*after the discussion, documentation of goals of care was present in the medical record*”

7) “*after the discussion, the goals of care present in the medical record are consistent with the patient’s stated preferences*”.

### Appendix 6. Evaluation of the online training program

Tables A1, A2, and A3 present the participating intensivists’ assessment of the different parts of the online training program, their level of self-confidence to use shared decision making and the context-adapted DA before and after the training program.

**Table A1 Overall assessment of the online training program by the participating intensivists (*n* = 5)**

| **What is your overall assessment of the following elements?** | **Low** | **Good** | **Excellent** |
| --- | --- | --- | --- |
| Organization and content (n, %) | 0 (0) | 1 (20) | 4 (80) |
| Clarity of information (n, %) | 0 (0) | 1 (20) | 4 (80) |
| Mastery of the subject by the creators (n, %) | 0 (0) | 2 (40) | 3 (60) |
| Clarity of explanations on the use of decision aids (n, %) | 0 (0) | 1 (20) | 4 (80) |
| Role play (n, %) | 0 (0) | 3 (60) | 2 (40) |
| General quality of the training (n, %) | 0 (0) | 1 (20) | 4 (80) |

**Table A2 Assessment of the online training program’s objectives by the participating intensivists (*n* = 5)**

| **Do you agree with the following statements?** | **Totally disagree** | **Disagree** | **Agree** | **Totally agree** |
| --- | --- | --- | --- | --- |
| Objectives of the training were clear and precise (n, %) | 0 (0) | 0 (0) | 2 (40) | 3 (60) |
| Objectives of the training were achieved (n, %) | 0 (0) | 0 (0) | 2 (40) | 3 (60) |
| Content of the training was adapted to my needs (n, %) | 0 (0) | 1 (20) | 2 (40) | 2 (40) |

**Table A3 Self-confidence to use shared-decision making and the context-adapted decision aid to support reported by the participating intensivists**

| **When supporting elders to make decisions about goals of care, how would you estimate your self-confidence level to use (…)? ^*^:** | **Before training program**  **(*n* = 5)** | **After training program**  **(*n* = 5)** |
| --- | --- | --- |
| Shared decision making, median score (%) [IQR^**^] | 8 [8-9] | 9 [9-9] |
| Context-adapted decision aid, median score (%) [IQR] | 7 [7-8] | 9 [9-9] |

^*^ Likert scale ranging from 0 to 10: 0 = not confident at all and 10 = totally confident)

^**^IQR: 25-75% interquartile range

### Appendix 7. Post-study feedback analysis of barriers and facilitators

### Please write to the corresponding author for original quotes in French.

| Barriers | Quotes  (*n*) | Representative quotes | Summary of findings |
| --- | --- | --- | --- |
| Physician attitude and behavior | **19** | ***I found it really difficult to go talk right away to them [the patients].***  ***I talk [about goals of care] only when the patients are 80, 85 years old…***  ***[…] we will surely have patients that it is not really necessary to talk to, sometimes you might scare them.***  ***All change requires a period of adaptation. Since we were not required or forced to use it [the DA], well, most of us didn’t use it.***  ***At the psychological level, to give a pamphlet to a patient about a very important subject like this, maybe I had the impression I was oversimplifying a complex question to make it mundane […]*** | Physicians believe that the discussion should be tailored to elderly patients, nevertheless they felt uncomfortable talking about goals of care with their patients. They were afraid of creating unnecessary anxiety and were afraid to turn a complex and sensitive issue into something commonplace or mundane. This takes time.  The physicians also discussed the difficulty to change their habits and practices in the ICU. Particularly for late adopters, there may be a lack of perceived benefits due to limited usage and experience with the DA. This takes time and may have to be imposed.  Similarly, there’s a concern about the fact that using a DA could cause anxiety in patients, particularly in short-term ICU stays or patients admitted after elective surgeries. |
| Reality of critical care practice | **16** | ***I wouldn’t give this [decision aid] without any explanation. Not to someone in a very acute condition.***  ***I had the feeling that I didn’t know them [the patients] enough, that I couldn’t take the time to discuss their goals of care, when my only goal was to see the patient and […] discharge them to a less acute ward [out of the ICU].***  ***So, I think, if we give this document to this patient, and that, the nurses, the residents, that nobody is able to answer the patient’s questions and that it falls back on us [the physicians] again… I have the impression, ... I am not certain that I will have the time to discuss this tool with the patient […].*** | ICU clinical situations and patients might not be ideal to start a delicate SDM discussion and to use the DA.  Indeed, many situations in this environment are complex, uncertain, need immediate attention, or are a source of anxiety for both the health professionals and the patients. The physicians have many tasks and frequently lack time to sit and discuss with the patients and don’t have the time or ability to answer their questions.  At the moment, only physicians are trained to conduct SDM and to use the DA. This adds extra pressure on them.  Patients may not be able to discuss and share their preferences. |
| Misunderstanding about SDM and the use of the DA | **6** | ***Participant: Not take a decision, but if you give the pamphlet in a community health and social service clinic […] Ah! It’s not the right tool…***  ***Interviewer: It’s a decision aid, not a pamphlet.***  ***We are trying to personalize our discussion in function of the pathology and of the patient, and this is somewhat complex. So, if you make one generic tool for all patients… it’s very limiting.*** | Physicians seemed to be confused about the goal of the DA, its content, and how and when it could be used in the ICU (e.g., before, during or after meeting a patient).  For instance, physicians seem to have understood that the recommended method for using the DA was to provide the tool to the patient without going into further explanations or discussions on the subject.  The length of the DA (ie, number of pages, number of words) and its design still needs to be worked on. The DA was originally designed with ICU patients who were probably not as ill as the ones in this implementation study. |
| Lack of practical training | **3** | ***From what I remember, […] I think that it [the training] was lacking something practical.*** | Their training about SDM and DA was also very theoretical and didn’t have enough concrete examples and practical applications on how to implement principles of SDM. Need for more video examples and practice.  Other ICU health professionals weren’t trained to engage in SDM and to use the DA. |
| Change management issues | **3** | ***To be honest, I didn’t know if I had the right to use it or not. No, but seriously, there was a time when we weren’t able to use it [the decision aid]. [...] And after I was all confused if I could use the decision aid or not.*** | The physicians were confused on when they could use the DA during the course of the study. |

| Facilitators | Quotes  (*n*) | Representative quotes | Summary of findings |
| --- | --- | --- | --- |
| Physician behaviors and attitude | **8** | ***I am really happy to know that I can use it.***  ***If we discuss with the patient and we explain, and we give him the level of care and he needs to be intubated. We need the tool (the DA). I think that if we go see the patient, and we give him the tool and he want to have time to reflect, we can give the handout.***  ***We need a human relationship with the patient.*** | Some physicians in the ICU were enthusiastic to use the decision aid and to have the discussion. The physicians also indicated many behaviors that could help them engage in goals of care discussions.  The physicians in the ICU want to personalize the discussion they have with their patients and want to take the time to develop a good human-centered relationship with their patients where their values and preferences are taken into consideration. They also suggest that informing the patient is important before discussing with them. |
| A simple and clear decision aid (DA) | **5** | ***I wouldn’t give this [decision aid] without any explanation. Not to someone in a very acute condition.*** | Physicians express their interest for a simple and clear tool that the patient could use by themselves.  They also appreciated the values clarification section because emphasis was on determining objectives of care and not only the solutions (CPR and mechanical ventilation). |
| Public and patient education before ICU | **4** | ***As an example, I have the pamphlet at home, I am not critically sick, I am not about to die. Well, I think I can look at that [the decision aid] relatively calmly.*** | The physicians suggested discussing goals of care with patients before they are hospitalized as a general public health approach.  This could give patients time to understand, to ask questions, and to receive the information in a calm environment.  Discussions about the goals of care should be done in primary care. |
| Training with emphasis on the evidence-based tools that will be used | **4** | ***I don’t know… It would be appropriate to incorporate this tool in the practice of all [health professionals]. So here’s what we can discuss with this [the decision aid]. Do you understand?*** | Physicians suggested training health professionals using a more practical than theoretical approach and then to force physicians to use it in the ICU (top-down mandated approach).  The training should also emphasize the fact that the DA was evidence-based and was previously patient-tested. |
| Family member inclusion | **2** | ***It would have been easier to give this tool [the decision aid] in the waiting room, and in the family room. They look at it, and then you come and talk to them about the levels of care. They look at it. You ask questions.*** | Physicians suggested to include family members in the discussions and to give them the decision aid too since many patients are sometimes unable to discuss their own levels of care. |
| Team support in the ICU | **2** | ***Let’s say that I am a patient and that I had that [decision aid], and that I have a question or that I want to talk, but the intensivist is not there. I mean, is there anyone else that could help if I have some important questions?*** | Physicians have identified a gap in the training of other healthcare professionals within the ICU, highlighting that aside from intensivists, there is a lack of proficiency in SDM and knowledge about how to use DAs.  To establish a more comprehensive and collaborative approach to DA use and goals of care discussions, they suggested extending training beyond intensivists, to ensure that all team members are well-versed in these skills.  This approach aims to reduce the reliance solely on intensivists and foster a team-based approach to goals of care discussions.  Surgeons should be trained to discuss goals of care discussions in the preoperative clinic for elective post-operative ICU admissions. In this setting, patients could consult the DA before being admitted to the ICU and have more time to reflect on what is important to them. |
| Patient attitudes | **1** | ***So, [the decision aid] could help us in this situation. We are always trying to put ourselves in the shoes of the patients… and if we feel like they accept this, well, it could [be used, …] if everybody feels comfortable.*** | The discussion about the goals of care can be facilitated if the physician feels that the patient is ready and accepts the situation. |

### Appendix 8: Standards for Reporting Implementation Studies: the StaRI Checklist

| **Checklist item** | | **Implementation strategy** | **Intervention** | **Page number or section** |
| --- | --- | --- | --- | --- |
| **Title** | **1** | Identification as an implementation study, and description of the methodology in the title and/or keywords | | Keywords, p.2 |
| **Abstract** | **2** | Identification as an implementation study, including a description of the implementation strategy to be tested, the evidence-based intervention being implemented, and defining the key implementation and health outcomes | | Abstract, methods section, p.1 |
| **Introduction** | **3** | Description of the problem, challenge, or deficiency in healthcare or public health that the intervention being implemented aims to address | | Introduction, second sentence, p.3 |
|  | **4** | The scientific background and rationale for the implementation strategy (including any underpinning theory, framework, or model, how it is expected to achieve its effects, and any pilot work) | The scientific background and rationale for the intervention being implemented (including evidence about its effectiveness and how it is expected to achieve its effects) | Introduction, first paragraph p.3 |
| **Aims and objectives** | **5** | The aims of the study, differentiating between implementation objectives and any intervention objectives | | Introduction, second paragraph p.3 |
| **Methods: description** | **6** | The design and key features of the evaluation (cross referencing to any appropriate methodology reporting standards) and any changes to study protocol, with reasons | | Methods, section 2.1 Design, p.3 and 4 |
|  | **7** | The context in which the intervention was implemented (consider social, economic, policy, healthcare, organisational barriers and facilitators that might influence implementation elsewhere) | | Methods, section 2.2, p. 4. See also Tables 1 and 2 on page 10. |
|  | **8** | The characteristics of the targeted “site(s)” (locations, personnel, resources, etc) for implementation and any eligibility criteria | The population targeted by the intervention and any eligibility criteria | Methods, section 2.2 and section 2.3 on p.4 |
|  | **9** | A description of the implementation strategy | A description of the intervention | Methods, section 2.4, p.4 and p.5 |
|  | **10** | Any subgroups recruited for additional research tasks, and/or nested studies are described | | Not applicable |
| **Methods: evaluation** | **11** | Defined pre-specified primary and other outcome(s) of the implementation strategy, and how they were assessed. Document any pre-determined targets | Defined pre-specified primary and other outcome(s) of the intervention (if assessed), and how they were assessed. Document any pre-determined targets | Methods, section 2.5.1; paragraph 3 on page 5 and; section 2.5.2, first sentence under “Outcomes”, p.5. |
|  | **12** | Process evaluation objectives and outcomes related to the mechanism(s) through which the strategy is expected to work | | Methods, section 2.5.5; first paragraph, p.7. |
|  | **13** | Methods for resource use, costs, economic outcomes, and analysis for the implementation strategy | Methods for resource use, costs, economic outcomes, and analysis for the intervention | Not applicable |
|  | **14** | Rationale for sample sizes (including sample size calculations, budgetary constraints, practical considerations, data saturation, as appropriate) | | Methods, section 2.6; first paragraph, p.7. |
|  | **15** | Methods of analysis (with reasons for that choice) | | Methods, section 2.7, Quantitative data analysis and qualitative data analysis, p.7 and p.8. |
|  | **16** | Any a priori subgroup analyses (such as between different sites in a multicentre study, different clinical or demographic populations) and subgroups recruited to specific nested research tasks | | Not applicable |
| **Results** | **17** | Proportion recruited and characteristics of the recipient population for the implementation strategy | Proportion recruited and characteristics (if appropriate) of the recipient population for the intervention | Results, section 3.1 on p.8, Table 1 on p.9, and Participant Flow chart on p.8. |
|  | **18** | Primary and other outcome(s) of the implementation strategy | Primary and other outcome(s) of the intervention (if assessed) | Results, section 3.2 on p.9 and p.10 |
|  | **19** | Process data related to the implementation strategy mapped to the mechanism by which the strategy is expected to work | | Results, section 3.7 on p.15 |
|  | **20** | Resource use, costs, economic outcomes, and analysis for the implementation strategy | Resource use, costs, economic outcomes, and analysis for the intervention | Not applicable |
|  | **21** | Representativeness and outcomes of subgroups including those recruited to specific research tasks | | Not applicable |
|  | **22** | Fidelity to implementation strategy as planned and adaptation to suit context and preferences | Fidelity to delivering the core components of intervention (where measured) | Results, section 3.2 on p.9 and p.10 |
|  | **23** | Contextual changes (if any) which may have affected outcomes | | Not applicable |
|  | **24** | All important harms or unintended effects in each group | | Results, sections 3.5 and 3.6 on p.13 to p.15, discussing concerns and misunderstandings |
| **Discussion** | **25** | Summary of findings, strengths and limitations, comparisons with other studies, conclusions and implications | | Discussion, p.16 to p. 19 |
|  | **26** | Discussion of policy, practice and/or research implications of the implementation strategy (specifically including scalability) | Discussion of policy, practice and/or research implications of the intervention (specifically including sustainability) | Discussion, p.19 |
| **General** | **27** | Include statement(s) on regulatory approvals (including, as appropriate, ethical approval, confidential use of routine data, governance approval), trial or study registration (availability of protocol), funding, and conflicts of interest | | Declarations, p.22 |

### Appendix 9: COnsolidated criteria for REporting Qualitative research (COREQ) Checklist

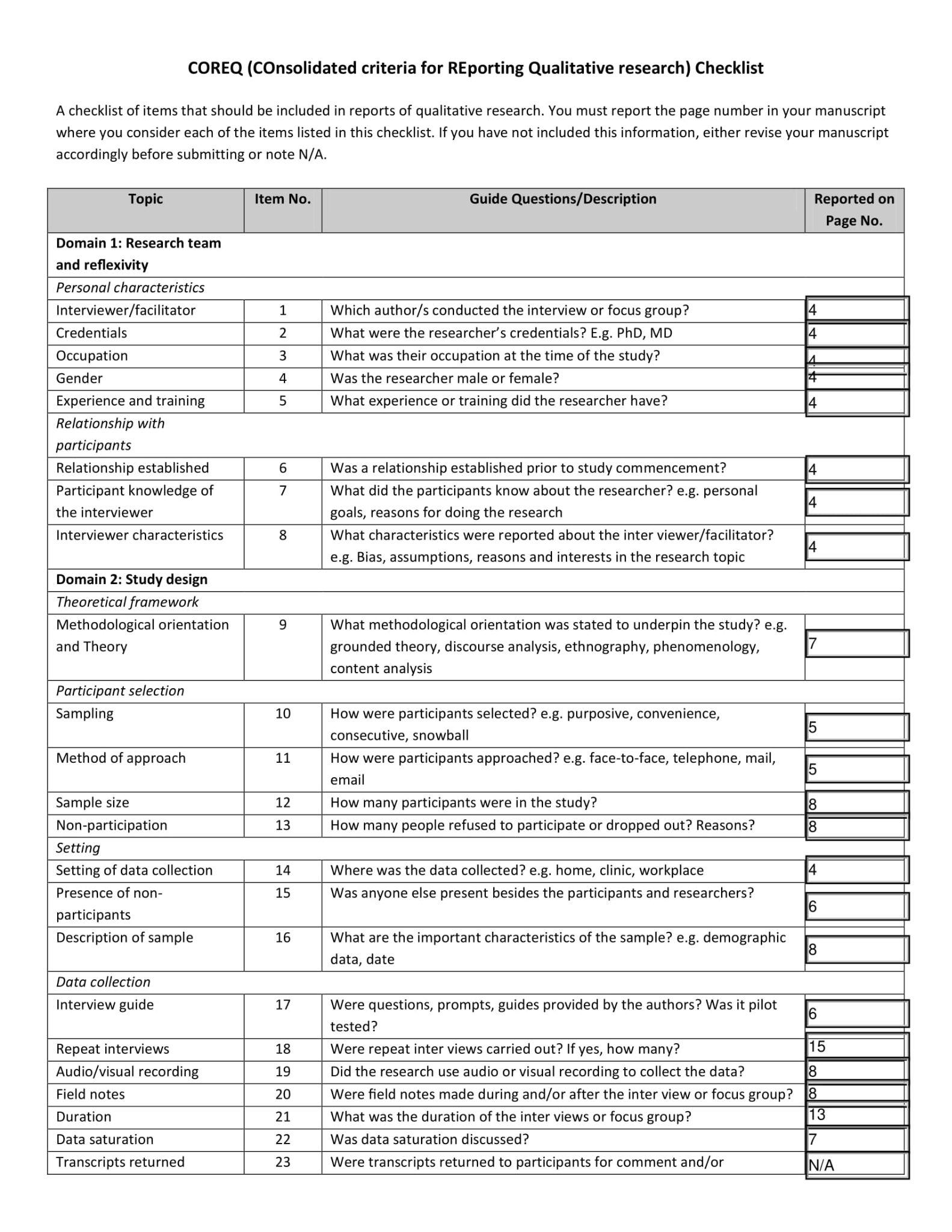

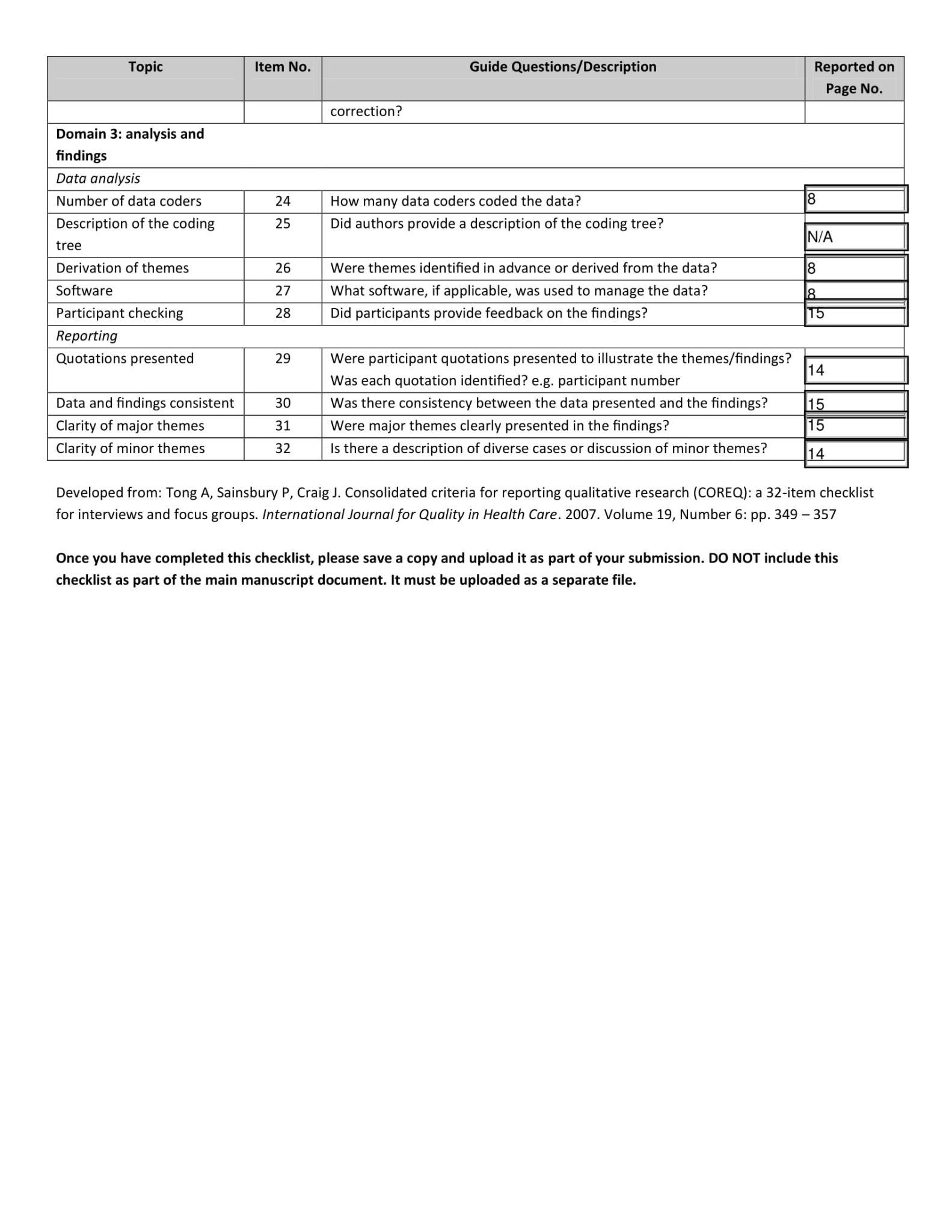

1. Sciences de la santé, Université du Québec à Rimouski (UQAR), Lévis, Québec, Canada
2. Institut universitaire de cardiologie et de pneumologie de Québec (IUCPQ), Department of anesthesia and critical care, Faculté de médecine, Université Laval, Québec, Canada
3. VITAM - Centre de recherche en santé durable, Québec, Québec, Canada
4. Centre de recherche en santé durable, Centre intégré universitaire de santé et de services sociaux de la Capitale-Nationale (CIUSSS-CN), Québec, Canada
5. Department of Family Medicine and Emergency Medicine, Faculté de médecine, Université Laval, Québec, Canada
6. Centre de recherche intégrée pour un système apprenant en santé et services sociaux, Centre intégré de santé et services sociaux de Chaudière-Appalaches (CISSS-CA), Lévis, Québec, Canada
7. Centre hospitalier universitaire de Québec-Université Laval (CHUL), Québec, Québec, Canada
8. Faculté de médecine, Université Laval, Québec, Québec, Canada
9. Université TÉLUQ, Québec, Québec, Canada
10. Centre de recherche et d’innovation en technologie éducative du Québec (i-TEQ)
11. Faculté des sciences infirmières, Université Laval, Québec, Québec, Canada
12. Centre de recherche hospitalier universitaire de Québec-Université Laval (CHUL; Oncology Axis), Québec, Québec, Canada
13. Center for Gerontology and Healthcare Research, Brown University School of Public Health, Providence, Rhode Island, USA
14. Department of Epidemiology and Biostatistics, College of Public Health, University of Georgia, Athens, Georgia, USA
15. 15 Research Group Technology for Health, HAN University of Applied Sciences, Nijmegen, The Netherlands
16. Centre de recherche hospitalier universitaire de Québec-Université Laval (CHUL; Population Health and Optimal Practices Unit), Québec, Québec, Canada
17. Department of Anesthesiology and Critical Care Medicine, Division of Critical Care Medicine, Faculté de médecine, Université Laval, Québec, Canada
